# Genomic investigation of a dengue virus outbreak in Thiès, Senegal, in 2018

**DOI:** 10.1101/2020.11.25.20235937

**Authors:** Amy Gaye, Tolla Ndiaye, Mouhamad Sy, Awa B. Deme, Alphonse B. Thiaw, Aita Sene, Cheikh Ndiaye, Younouss Diedhiou, Amadou M. Mbaye, Ibrahima Ndiaye, Christopher Tomkins-Tinch, Jules F. Gomis, Aida S. Badiane, Bronwyn MacInnis, Daniel J. Park, Mouhamadou Ndiaye, Ngayo Sy, Pardis C. Sabeti, Katherine J. Siddle, Daouda Ndiaye

**Author notes:** These authors contributed equally to this article (Amy Gaye, Tolla Ndiaye, Mouhamad Sy). These authors contributed equally to this article (Katherine J. Siddle, Daouda Ndiaye). Corresponding author: Daouda Ndiaye, Address for correspondence: Université Cheikh Anta Diop, Dakar, Senegal, BP 16477 | |.

## Abstract

Dengue virus is a major and rapidly growing public health concern in tropic and subtropic regions across the globe. In late 2018, Senegal experienced its largest dengue virus outbreak to date, covering several regions. However, little is known about the genetic diversity of dengue virus (DENV) in Senegal. Here we used molecular tools including metagenomic sequencing to identify 19 previously undetected dengue virus cases from the city of Thiès and assemble 17 complete virus genomes. DENV3 was the most frequent serotype; 11 sequences (65%) were DENV3, 4 sequences were DENV2 and 2 were DENV1. Sequences were closest to recent sequences from West Africa, suggesting ongoing local circulation of viral populations; however, detailed inference is limited by the scarcity of available genomic data. We did not find clear associations with reported clinical signs or symptoms, highlighting the importance of testing for diagnosing febrile diseases. Overall, these findings expand the known range of DENV in Senegal, and underscore the need for better genomic characterization of DENV in West Africa.

## Introduction

Dengue virus is a major and growing problem across the world, with an estimated 390 million infections per year ^1^. Recent years have seen large outbreaks in endemic areas across Asia, Africa and the Americas as well as the appearance of the disease in new areas. Climatic and demographic changes, including urbanization and increasing temperatures, may be contributing to this emergence, creating more favourable conditions for the Aedes mosquitoes, the disease vector ^2,3^. The majority of dengue virus infections are either mild or asymptomatic; these can be difficult to diagnose as symptoms - commonly including fever, myalgia, arthralgia, and headaches - are usually non-specific, meaning diagnostic tests are needed to confirm infection. A small fraction of cases result in a severe disease, especially following secondary infection with a different serotype of the virus via antibody-dependent enhancement ^4^.

Dengue is endemic to 34 countries across Africa, and outbreaks of this disease have been reported in all regions ^5^. Dengue is caused by a virus of the Flaviviridae family and there are 4 distinct serotypes (DENV1, DENV2, DENV3 and DENV4). All four serotypes have been reported in Africa, with DENV2 being associated with the greatest number of epidemics on the continent ^5^. In Senegal, dengue virus was first isolated in 1970 in the rural Bandia area ^6^. All serotypes have since been detected in Senegal, with DENV3 causing an epidemic in 2009 Dakar ^7^ and DENV2, often confined to the forest cycle, spilling over into urban areas in Senegal in 2014–2015 ^8^. Since then, recurrent urban outbreaks associated with multiple serotypes have been reported in both 2017 (138 cases) and 2018 (314 cases) ^8,9^, raising the possibility that environmental changes and high mobility of people has led to a more widespread distribution of dengue virus across the country.

In late 2018, Senegal experienced the largest dengue outbreak to date, with 314 confirmed cases detected in several regions of the country. Over 60% of cases were in the region of Diourbel, concentrated in the city of Touba, where in late October the Grand Magal -- a 2-day religious pilgrimage that gathers over 3 million people in this holy city -- took place ^10,11^. Given the risk of disease transmission at this large gathering, additional measures to enhance disease surveillance were put in place, including a mobile laboratory from the Institut Pasteur equipped with molecular tests for rapid diagnosis ^10^. However, several factors indicate that many cases were undetected: the incubation period of the disease is typically 5-7 days, dengue fever was detected elsewhere in Senegal even before the Magal, and the similarity in symptoms and seasonality of dengue fever and malaria suggest that many more cases, especially of mild disease, remained undetected. Dengue is, therefore, likely to be an underdiagnosed cause of non-malarial fever.

Genomics can provide important information regarding the origin, evolution and spread of a viral outbreak, such as dengue ^12–14^. Here we used metagenomic sequencing to characterize the genetic diversity of a cluster of previously undetected dengue virus cases from the city of Thiès between October and November 2018, and we evaluated clinical metadata to identify risk factors associated with dengue virus infection. Despite the growing burden of dengue fever in Senegal and West Africa more broadly, little is known about the genetic diversity of the virus and few viral genomes are available. This information could help explain disease transmission dynamics and enable a better understanding of dengue virus circulation and transmission in Senegal.

## Methods

### Sample collection

Patient samples were obtained through a study approved by the Institutional Review Boards at the Senegalese Ministry of Health and Harvard University. Patient samples were obtained from individuals presenting to the Section de Lutte AntiParasitaire in Thiès, Senegal. Study staff obtained written, informed consent from all patients enrolled in the study, following an explanation of the study objectives. A 5mL venous blood draw was collected from each enrolled patient in a BD Vacutainer EDTA tube. Demographic information as well as clinical signs and symptoms were also recorded. Blood samples were kept on ice and transported daily to the Laboratoire Parasitologie et Mycologie at Aristide Le Dantec Hospital, where plasma was isolated on the same day and frozen prior to RNA extraction.

### RNA Extraction and PCR

Total RNA was extracted from patient plasma using the QiAmp viral RNA mini kit (Qiagen) according to the manufacturer’s instructions. Detection of Flavivirus RNA was performed by RT-qPCR using Applied Biosystems® Power SYBR® Green on a Roche LightCycler 96 for 45 cycles. Primer sequences have been shown to capture all known flavivirus species ^15^. Qualitative detection of Dengue virus RNA in samples with suspected flavivirus infections was performed using the RealStar® Dengue RT-PCR Kit 3.0 from Altona-Diagnostics on an Applied Biosystems® 7500 fast real-time PCR machine according to manufacturer’s instructions.

### Sequencing and data analysis

Sequencing libraries were prepared from all dengue virus positive samples and 17 negative samples. Briefly, DNA was removed by Turbo DNase treatment, cDNA was synthesized with random primers and sequencing libraries were prepared using the Nextera XT kit (Illumina) as previously described ^16^. All samples were sequenced on an Illumina MiSeq instrument at the Laboratoire Parasitologie and Mycologie in Dakar, Senegal, with paired end 100nt sequencing.

Sequencing data was analysed using viral-ngs v1.25.0 ^17^, implemented on the DNAnexus cloud platform. First, samples were demultiplexed and an initial taxonomic classification of metagenomic sequencing reads was performed using KrakenUniq ^18^. Reads were compared against a database that encompassed the known diversity of all viruses that infect humans. Next, sequences were filtered by removing reads aligning to the human genome, synthetic spike-in sequences and other known contaminants. The remaining reads were filtered against a database of dengue virus sequences, and *de novo* assembly was performed with Trinity ^19^. Contigs were then scaffolded against one of four RefSeq genomes, representing the four serotypes (NC_001477.1, NC_001474.2, NC_001475.2, NC_002640.1). Short read sequence data and assembled genomes have been deposited in NCBI’s SRA and Genbank databases under BioProject PRJNA662334 and accession numbers MW288024 - MW288040.

#### Code availability

Computational workflows used to analyze viral sequencing data are have been made publicly available through dockstore: https://dockstore.org/organizations/BroadInstitute/collections/pgs

### Phylogenetic tree construction

Genotype Detective (https://www.genomedetective.com/app/typingtool/dengue/) was used to determine the genotype and serotype of the newly assembled dengue virus genomes, as well as the genotypes of all whole dengue virus genomes (>10,000nt) available on GenBank on February 10th 2020 (DENV2 and DENV3) or June 24th 2020 (DENV1). We further filtered genome records to remove those where collection date or country of origin were not recorded. Due to the large number of non-African dengue genomes, a subset of these sequences was selected for phylogenetic analysis. Specifically, for each serotype, we randomly selected 50 published genomes from the same genotype as the current samples as well as up to 25 genomes from each other genotype for context. All complete sequences from Africa were also included (DENV1; N=10, DENV2; N=33, DENV3; N=7) yielding a final dataset of 136 genomes for DENV1, 188 genomes for DENV2 and 118 for DENV3.

Sequences were aligned using MAFFT v7.388 implemented in Geneious and trimmed with TrimAl. Maximum likelihood phylogenies were estimated separately for each serotype using IQTREE v1.5.5.5 ^20^, using a (GTR) + Gamma substitution model and ultrafast bootstrapping (N=1000) ^21^. Time-scaled Bayesian phylogenetic analysis was performed using a Markov chain Monte Carlo (MCMC) algorithm implemented in the BEAST v1.10.4 ^22^ on the CIPRES Science Gateway ^23^. Briefly, BEAST was run for 50 million MCMC steps using a GTR + Gamma substitution model with 1,2,3 codon partitioning, an uncorrelated relaxed clock with log-normal distribution and a Skyline tree prior. For each serotype we used the full coding sequence and sampled tip dates. Trees were sampled every 5,000 generations. The convergence was assessed using Tracer v1.7.1 and TreeAnnotator v1.10.4 was used to generate maximum clade credibility trees with a burn-in of 10%. All trees were viewed in FigTree v1.4.4 ^24^.

### Demographic and clinical data analysis

A univariate correlation analysis was performed on 14 categorical and 6 continuous variables to determine the significance of association with a presumptive diagnosis of dengue fever (determined by the presence of dengue virus RNA in the plasma detectable by RT-qPCR or sequencing). Analyses were performed as previously described ^25^. Briefly, the biserial correlation coefficient and the two-tailed Fisher Exact test were used to calculate the significance of continuous and categorical variables, respectively at a p-value cut-off of 0.05. Categorical variables tested were; sex, recent travel and the presence of 12 clinical signs or symptoms reported by at least 1 participant; muscle aches, diziness, vomitting, weakness, headache, backache, chest pain, difficulty breathing, rash, sore throat, inflammation and bleeding. Continuous variables tested were; age, temperature, heart rate, respiratory rate, hemoglobin level and blood sugar level.

## Results

### Study participation and location

Thiès is the third largest city in Senegal and the administrative capital of the region of Thiès. It is located 72km east of Dakar, Senegal’s capital and largest city, and 123km west of Touba, Senegal’s second largest city (Fig. 1A). Thiès has a very low malaria endemicity with an annual entomological inoculation rate of 1-5 ^26^. Individuals enrolled in this study presented to the SLAP clinic (Thiès) with either an acute fever (temperature >38C) or a recent history of fever in the last 4 days. All samples were collected between October 2nd and November 12th, a period just following the rainy season in this area of Senegal and when malaria transmission is at its annual peak ^27^. In total, 198 individuals were enrolled into the study, of whom 36 tested positive for malaria by Rapid Diagnostic Test. Individuals were aged between 2 and 72 years of age with 121 males and 77 females.

**Figure 1.**
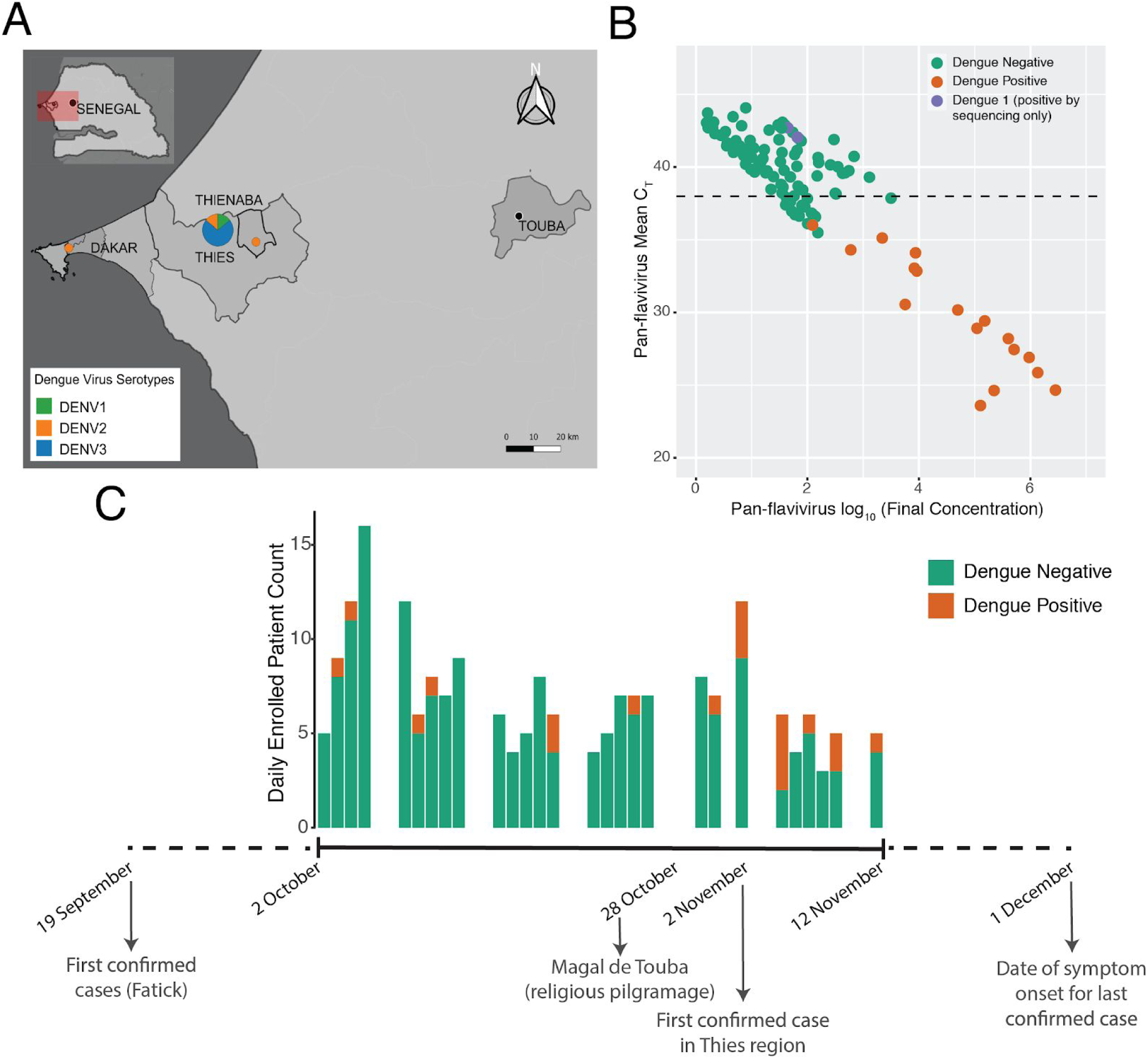
A. Map showing the location of Thiès, Senegal. B. Plot showing the CT and viral concentration of all 198 samples tested. Dengue-positive samples, as determined by the RealStar assay, are coloured in orange. C. Timeline of the 2018 dengue virus outbreak in Senegal showing the number of patients enrolled per day and the number of individuals that tested positive for dengue. Below the plot are marked several important events during the outbreak.

### Molecular detection of dengue virus among febrile cases

Following RNA extraction, all 198 samples were tested for the presence of flavivirus RNA using an in house RT-qPCR assay with previously published primers (see Methods). 38 samples that were suggestive of viral presence were further tested using the RealStar® Dengue RT-PCR assay. This approach identified 17 positive samples (Fig. 1B). Coinfections of malaria and arboviruses have previously been reported in other regions of Senegal ^28^. Here, no samples that tested positive for malaria (N=36) also tested positive for dengue virus infection, suggesting no co-infections were present in our dataset. Dengue virus was detected in 9.6% of all febrile patients and 11.7% of all cases of non-malarial fevers.

### Metagenomic sequencing of dengue virus genomes

Using metagenomic sequencing we assembled complete viral genomes from 15 of the 17 positive samples (1 sample was excluded due to a low concentration of input material and another did not yield sufficient reads to assemble a high quality genome). The average coverage ranged from 28x to 5370x and the genome assembly length varied between 10,525 bp and 10,680bp (Table S1). Four samples were from serotype DENV2 and 11 samples were from serotype DENV3. No other pathogenic virus was detected in any of the samples sequenced (Fig. S1).

We also performed metagenomic sequencing on 17 samples that tested negative by the pan-flavivirus RT-qPCR assay. In 2 of these samples we detected sufficient reads to assemble complete genomes for DENV1, indicating that these were missed by the RT-qPCR assay. Indeed, sequence comparison of these 2 newly assembled genomes identified 4 mutations in the forward primer, likely impacting the performance of this assay (Fig. S2). We confirmed this result using the RealStar® Dengue RT-PCR. We also re-tested all available samples using a third RT-qPCR assay for Dengue virus that was able to detect these two positive samples and did not identify any other samples that were positive for DENV1.

### Phylogenetics of dengue virus in Senegal

We next investigated the phylogenetic relationships of these sequences to existing dengue virus genomes. DENV3 was the most frequent serotype in our data. All 11 genomes were highly similar and fell within an African cluster of samples in genotype III that also includes 5 sequences from Gabon and 1 from Senegal collected between 2009 and 2017 (Fig. 2B). However, the estimated TMRCA of these sequences with the most closely related sequences - from the Gabonese outbreak in 2016-17 - was ∼13 years ago (95% HPD 11.3 - 14.3 years; Fig. S3), suggesting that much of the recent evolutionary history of DENV3 is absent from our tree. To increase resolution to infer the recent history DENV3 in West Africa we repeated our phylogenetic analyses using a 935nt fragment of the envelope gene. This allowed the inclusion of 15 additional sequences from Africa including Cote d’Ivoire, and Cap Verde. These sequences were all part of the African genotype III cluster, but shared more similarity with the Gabonese and older Senegal sequence than the present sequences (Fig. S4).

**Figure 2.**
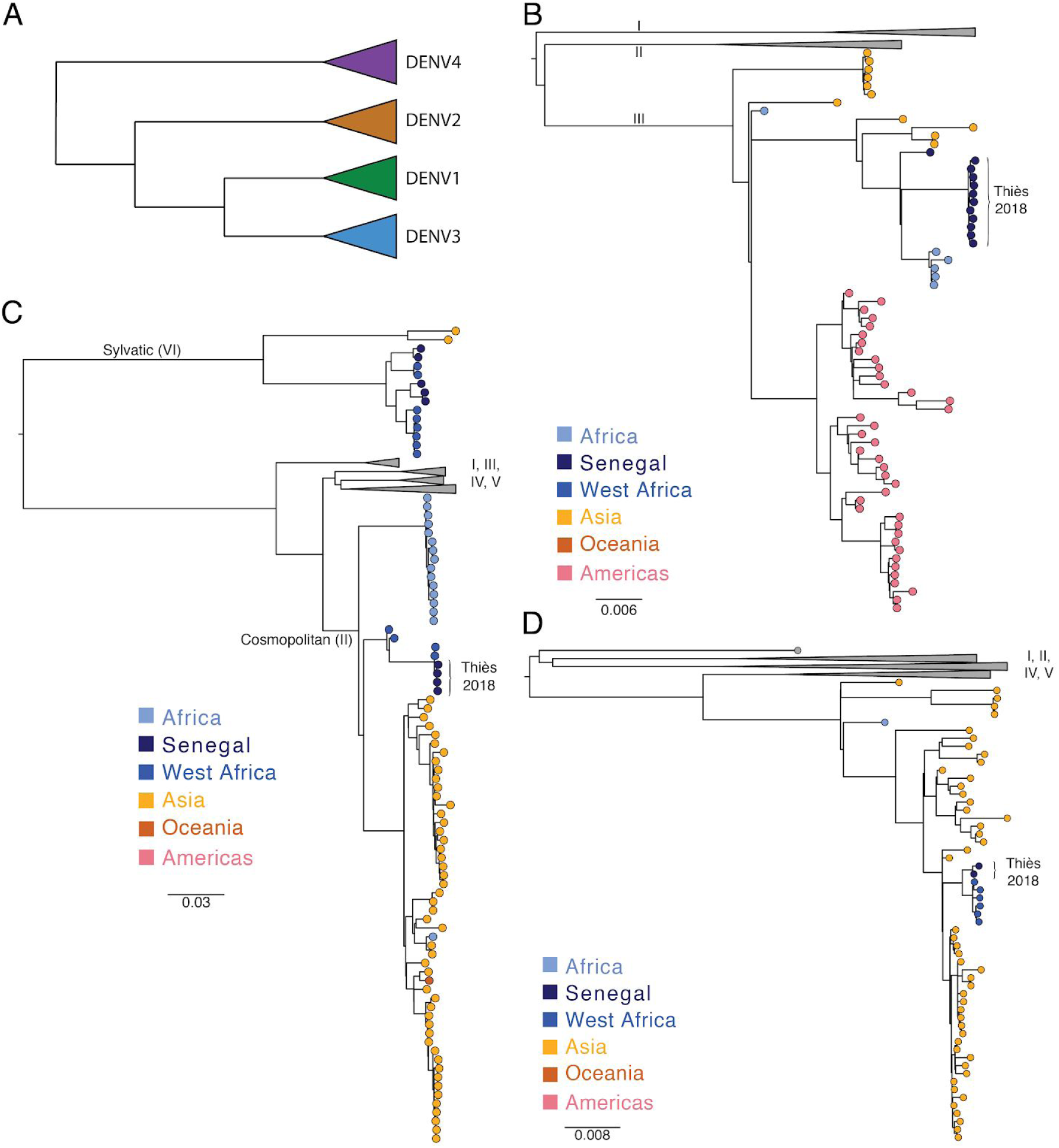
Phylogenetic analysis of dengue virus genomes from Senegal. A. Schematic of phylogeny of dengue virus serotypes. B. DENV2 maximum likelihood (ML) phylogeny: genomes from Thiès are in the Cosmopolitan genotype and are most similar to other West African genomes. C. DENV3 ML phylogeny: genomes from Thiès are in genotype III and are most similar to genomes from Africa. C. DENV1 ML phylogeny: genomes from Thiès are in genotype III and are most similar to genomes from Africa.

For DENV2, the four genomes all belonged to the Cosmopolitan genotype (II). Notably, these samples are not part of the sylvatic clade, unlike all previous DENV2 sequences that are available from Senegal (Fig. 2), consistent with the urban nature of this outbreak. All the new DENV2 genomes are highly similar and are closest to other available sequences from Africa, particularly those from Burkina Faso in 2016 (Fig. 2C). We estimated the time to the most recent common ancestor (TMRCA) of the Senegalese and Burkina Faso sequences to be 4 years), indicating a recent shared ancestor (Fig. S3). It is notable that Burkina Faso is the only other West African country from which complete genomes of the Cosmopolitan genotype are available. A shorter sequence from Cote d’Ivoire in 2017 (LC310791) also clustered with these sequences in an ML tree of only the envelope gene.

The two DENV1 genomes detected by metagenomic sequencing were from genotype III and most similar to the only other African complete genomes in this genotype (1 from Benin and 5 from Burkina Faso; Fig. 2D) with a TMRCA of ∼5 years ago (95% HPD 3.9 - 6.1 years; Fig. S3). Notably, these sequences cluster distinctly from the only other DENV1 sequences from Senegal - two short fragments of the polymerase gene from the Dakar suburb of Medina Gounass collected in 2015 ^29^, which cluster with Asian sequences of this genotype (Fig. S5). However, the short length of this sequence prevents more detailed phylogenetic relationships from being resolved.

### Association with clinical, demographic and epidemiological characteristics

Participants with presumptive dengue fever were of all ages (8-49; mean 27.79 years) with an equal number of males and females (Table 1). No clinical signs or symptoms were found to be differentially reported between presumptive dengue cases and dengue-negative febrile individuals. Headaches were the most common sign reported by all participants (197/198 participants including all presumptive dengue cases). Among participants with presumptive dengue fever, 89% (17/19) reported muscle aches and 79% (15/19) reported back aches. However, neither was significantly more common among these individuals, even after excluding individuals with a known malaria diagnosis (N=36) (Table 1).

**Table 1.**
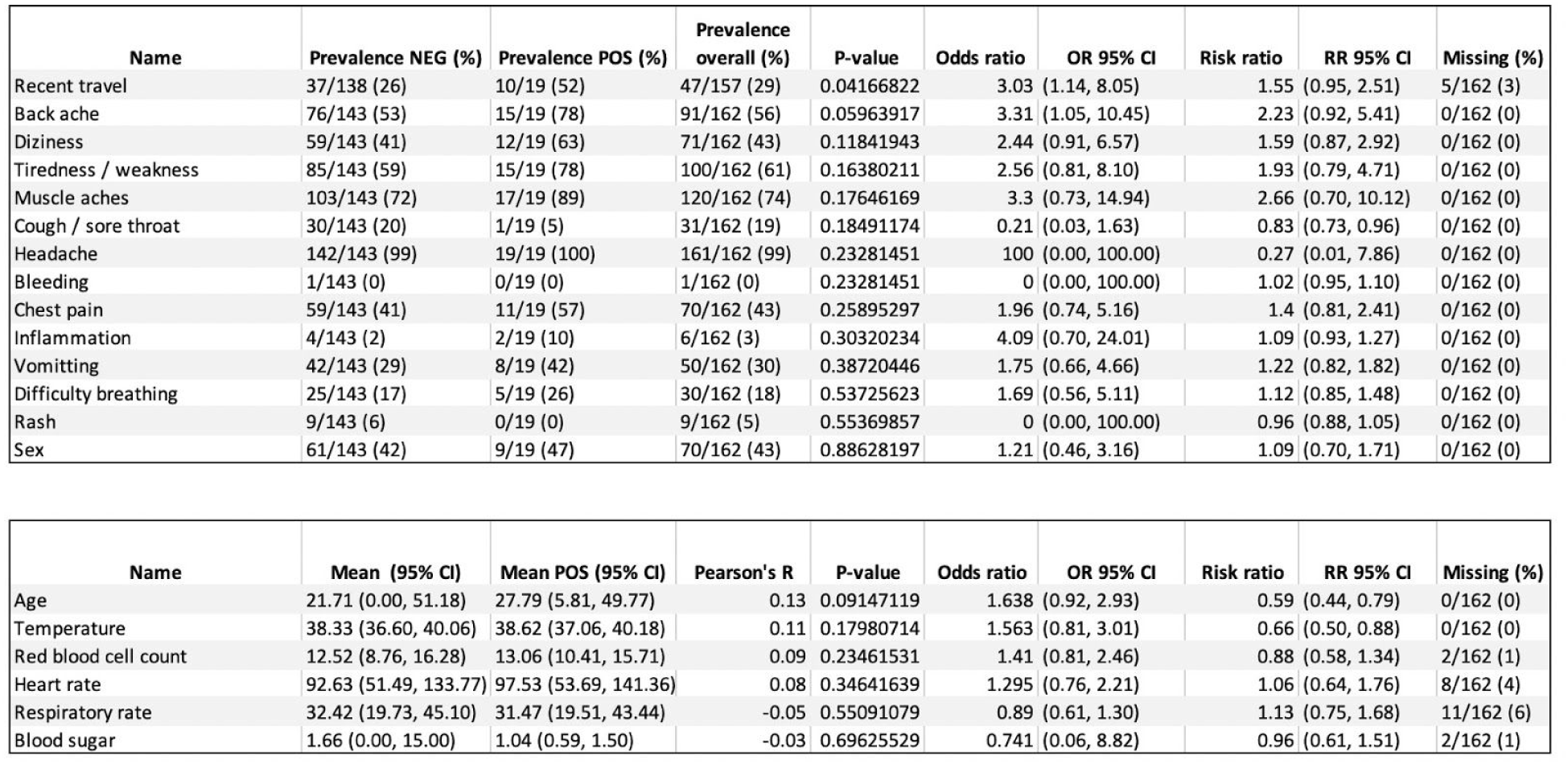
Association values for clinical and demographic variables. Top: binary variables and bottom: continuous variables.

Recent travel was significantly more frequent among individuals with presumptive dengue fever (52% vs 26%, p=0.041, Table 1). Nine of the ten individuals with presumptive dengue who reported recent travel had traveled to Touba (the other individual had traveled to Saint Louis). However, only 6 of these were after the dates of the Magal. All 6 individuals reported onset of fever within 10 days of the end of the Magal (the earliest on October 30th, the latest on November 6th). This timing could be consistent with the dates of the Magal, however, we do not know if the individuals attended the Magal and cannot rule out that they were infected elsewhere.

## Discussion

In this study we identify and describe the genomic diversity of a cluster of previously undetected dengue virus cases from the city of Thiès, Senegal. We report 17 complete DENV genomes (of serotypes 1, 2 and 3). This data enables the investigation of risk factors associated with dengue virus infection and is useful for the surveillance of repetitive outbreaks recorded in Senegal, where, as in other regions, urban outbreaks of multiple serotypes are increasingly common. These findings expand the known range of local transmission of dengue virus in late 2018 in Senegal and point to a much wider distribution of three serotypes of the virus than previously reported.

During the 2018 outbreak DENV1 and DENV2 were reported respectively from the southwest (Fatick) and north of Senegal (Saint Louis and Richard Toll); however, these findings suggest that it was more widespread. Of the 2 individuals with DENV1 infection, 1 reported travel to Touba, and the other reported no recent travel. Of the 4 individuals with DENV2 infection, 1 reported travel to Saint Louis prior to developing symptoms, 1 reported travel to Touba and the other 2 reported no travel, suggesting that DENV1 and DENV2 were circulating in the Thiès region. All four DENV2 isolates were genetically very similar and fell into the cosmopolitan (II) genotype, distinct from the 5 other genomes available from Senegal that are part of the sylvatic clade. The cosmopolitan genotype is widespread across Africa and Asia; however the present sequences are most similar to recent sequences from Burkina Faso in 2016. This suggests that the lineage of DENV2 associated with this outbreak was related to viruses that have been circulating in the West African region in recent years, however, the scarcity of sequence data prevents higher resolution on this.

The majority of cases in this study (65%) were from serotype 3. This serotype has become increasingly common in West and Central Africa in the last decade and has been responsible for several outbreaks in Senegal ^7^, Côte d’Ivoire ^5^, Gabon ^30^ and elsewhere. Our phylogenetic analysis suggests that this is a closely related but distinct lineage to that responsible for the 2009 outbreak in Senegal, as the samples cluster more closely to those from a 2016-17 outbreak in Gabon ^30^ than the single sequence from Senegal in 2009, sequenced from a traveler returning to Germany ^31^. However, we lack genomic data from this region; only 7 complete DENV3 genomes from Africa were available in GenBank prior to this study, in addition to 24 short sequences of 220 - 1,923nt. This scarcity of data makes phylogenetic inference, especially around the recent evolutionary history of DENV3 in West Africa, challenging.

In Senegal, the DENV3 serotype was most frequent in Touba (Diourbel) during the 2018 outbreak and the timing of the outbreak overlapped with the Grand Magal religious pilgrimage. 45% of DENV3 positive patients reported travel to Touba (N=5), of which 3 developed a fever in a time frame consistent with attendance at the Magal. It should be noted that we do not know when participants travelled to Touba or whether they attended the Magal. The other 6 individuals reported no recent travel. Even if we postulate that all individuals with recent travel were infected outside of Thiès, almost half of cases are consistent with locally-acquired infection.

Despite using a pan-flavivirus RT-qPCR for screening of all samples prior to sequencing, the two DENV1 infections were missed, owing to 4 mismatches in the forward primer binding region that compromised the sensitivity of the test. This finding demonstrates the utility of genomic surveillance, especially metagenomic approaches ^32^ for finding genetically distinct strains that may be missed by PCR-based approaches. It further emphasizes the importance of assembling and sharing complete viral genomes from all areas where dengue virus is endemic or emerging to ensure that targeted diagnostics, such as PCR primers, are sensitive to the full spectrum of genetic diversity circulating in the place and time of interest. Developing local capacity for this work in endemic countries is central to this effort and to ensuring pathogen genomics can be translated into actionable insights.

We found very little association between the clinical and demographic variables investigated and presumptive dengue virus infection. Individuals with presumptive dengue were equally distributed with regards to sex and age. Associations were stronger after removing patients with a confirmed malaria diagnosis, but still did not reach statistical significance. These findings are consistent with the mild symptoms of these individuals and the broader difficulty of clinically differentiating dengue fever from other febrile diseases without diagnostic testing. Although dengue fever does not have a specific therapy and the majority of infections are mild, diagnosis is still important to rule out other causes that would have different courses of treatment. The availability of rapid, sensitive and widely available diagnostic tests for common pathogens, other than malaria, is still severely limited in Senegal and many other regions and presents a major challenge for public health.

In conclusion, we report the analysis of 17 complete genomes from 19 previously undiagnosed dengue cases from the 2018 outbreak in Senegal. This work expands our understanding of the source and spread of this outbreak but also of dengue virus genetic diversity in West Africa more broadly. Our findings highlight the need for enhanced diagnosis and genomic surveillance to better understand the past and track the future course of this disease.

## Data Availability

Short read sequence data and assembled genomes have been deposited in the NCBI SRA and Genbank databases under BioProject PRJNA662334 and accession numbers MW288024 - MW288040.

## Acknowledgements

We thank the participants of this study and the staff of the Service de Lutte Antiparasitaire for their contributions to this work. We thank Andres Colubri for sharing code for the bivariate analyses, and Chris Edwards and Monica Schreiber for review of the manuscript. This work is supported by grants from the National Institute of Allergy and Infectious Diseases; NIH-H3Africa (U54HG007480) and the West African Emerging Infectious Diseases Research Center (U01 AI151812). K.J.S. is supported by the ASTMH Centennial Award. P.C.S. is an investigator supported by the Howard Hughes Medical Institute (HHMI).

## Author Contributions

A.B.D., A.S.B., B.L.M., D.J.P., M.N., N.S., P.C.S., K.J.S., D.N. conceived and designed the study; A.G., T.N., M.S., A.T., A.S., C.N., Y.D., A.M., I.N. acquired the data; A.G., T.N., M.S., A.B.D., J.G., K.J.S. analyzed the data; D.P., C.T-T. created software used in this work; A.G., T.N., M.S., A.B.D., K.J.S. drafted and revised the manuscript with input from all authors. All authors approved the manuscript before submission.

