## Supplemental Material for "Genomic investigation of a dengue virus outbreak in Thiès, Senegal, in 2018"

**Figure S1.** Heatmap of normalized counts for viral taxa from the metagenomic sequencing data using KrakenUniq. Shown are the viral species known to infect humans for which at least 1 of the 18 sequenced samples had >100 reads per million sequenced reads assigned to that species. Bacteriophages have been removed. Kyasanur forest disease virus, Powassan virus and Sepik virus are all flaviviruses, suggesting these may be misclassifications of reads within this large viral genus. Human associated gemykibivirus 2 is a virus of unknown pathogenic effect in humans.

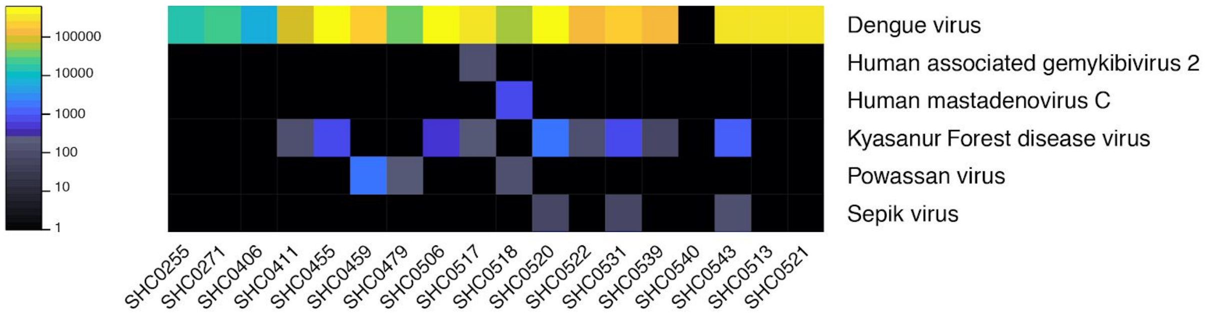

**Figure S2.** Alignment of select dengue virus genomes to pan-flavivirus primer. DENV1 sequences from Senegal (SHC0513 and SHC0521) showed presence of 4 mismatches (C8941T, C8950T, G8953A and G8962A) in the primer binding region of pan-flavivirus forward primer (in blue). This likely explains why these sequences were not detected by this assay, unlike DENV2 and DENV3.

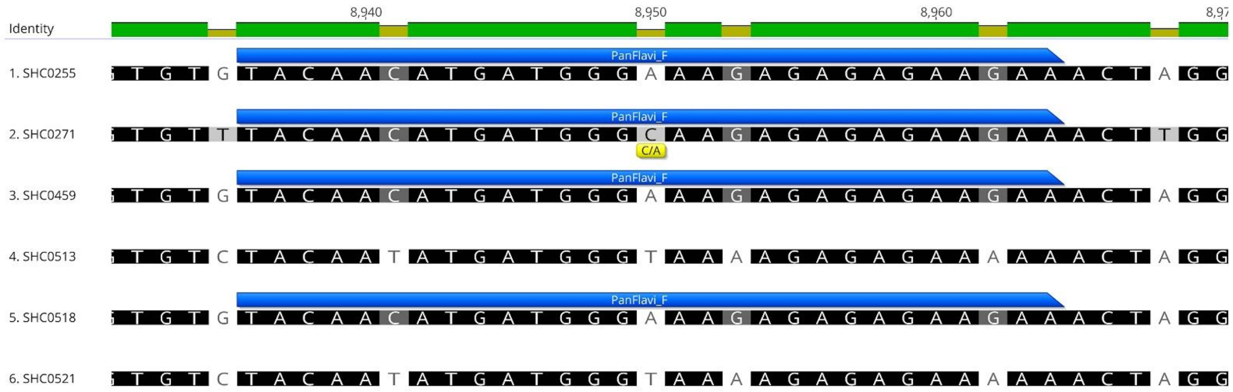

**Figure S3:** Maximum clade credibility trees for (A) DENV1 genotype III, (B) DENV2 Cosmopolitan genotype and (B) DENV3 genotype III. Estimated date and 95% posterior densities are given for nodes of interest described in the text.

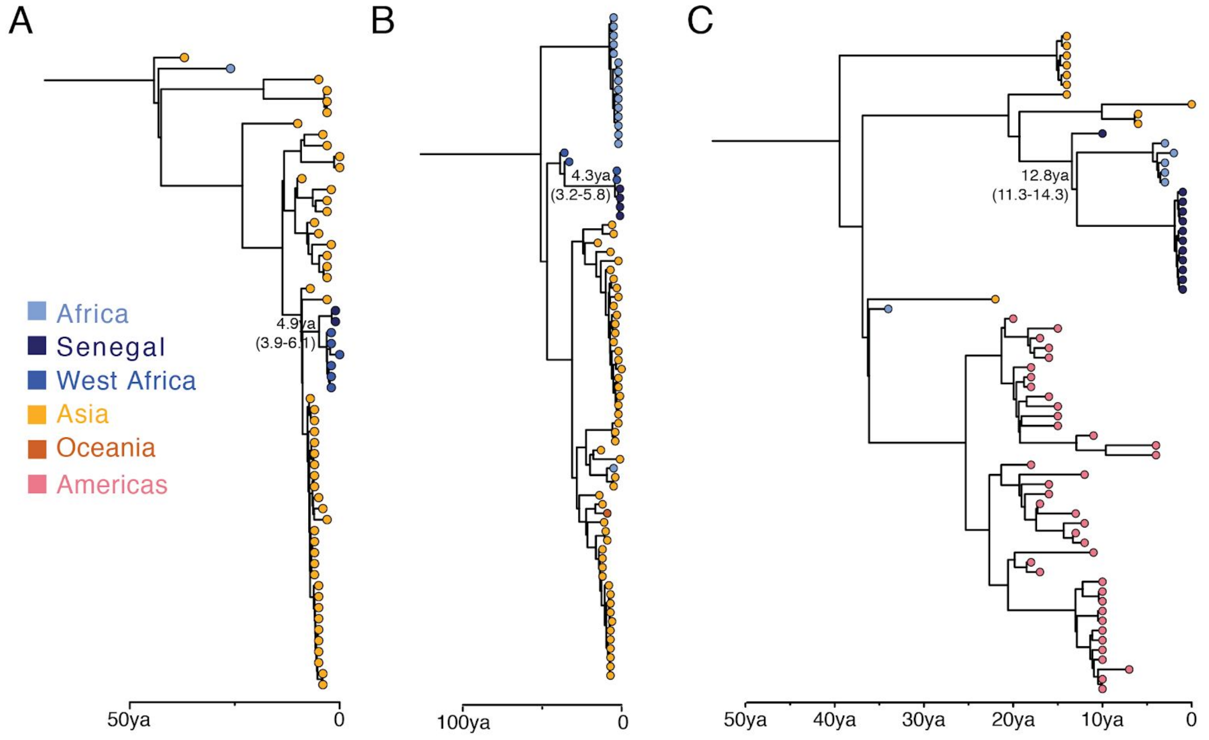

**Figure S4.** Maximum Likelihood tree of a 955nt fragment of the envelope gene of DENV3 showing the relationship between available African sequences. Tips are coloured by country and GenBank IDs are given for  available African sequences. All non-African sequences are gray. Bootstrap values are indicated on major nodes associated with African sequences.

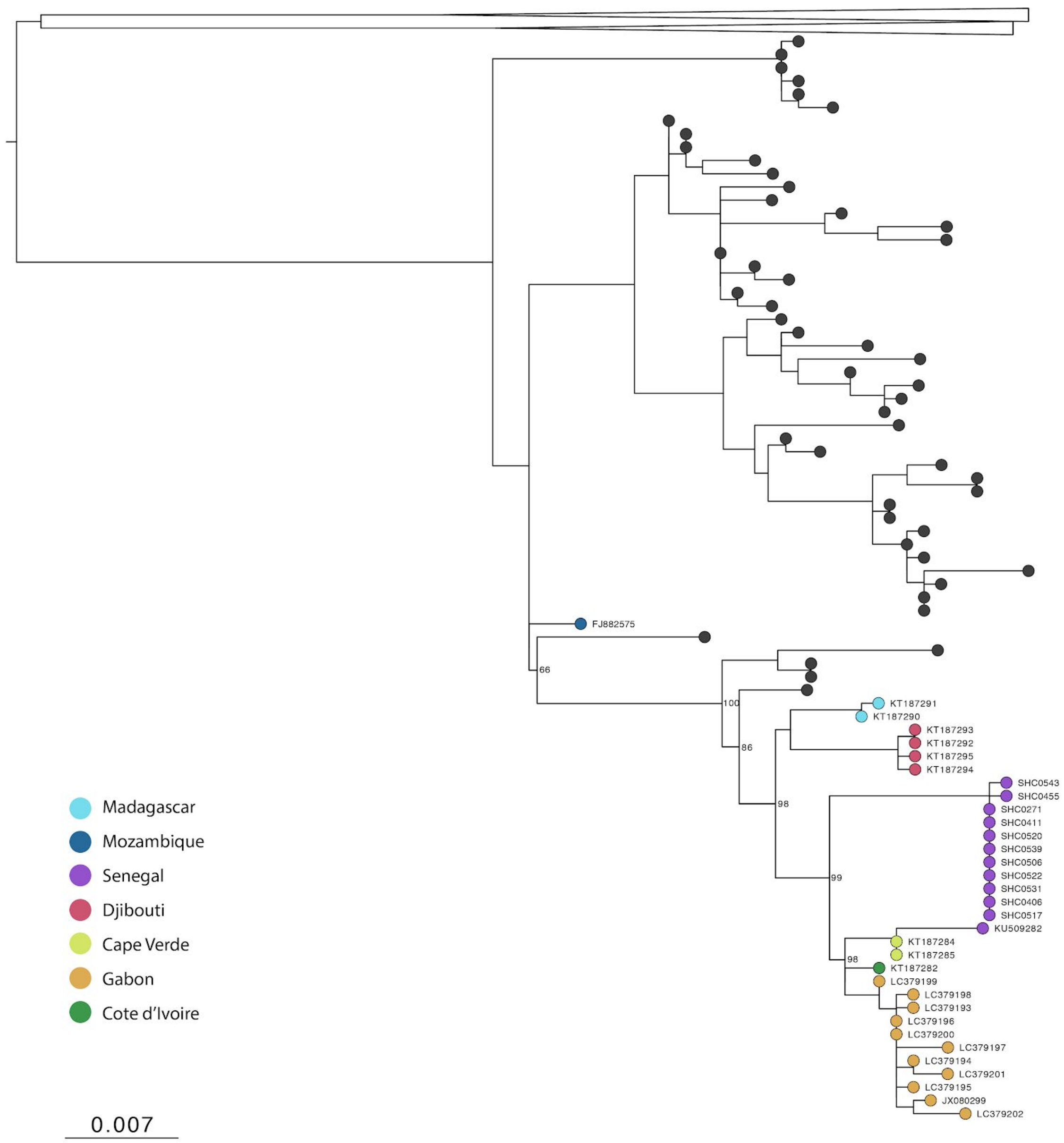

**Figure S5.** Maximum Likelihood tree of a 245nt fragment of the polymerase gene for 60 sequences from DENV1 genotype III. A single randomly selected sequence from each other genotype (I, II, IV and V) are included as outgroups. The new sequences from Senegal (SHC0521 and SHC0513) cluster with other sequences from elsewhere in West Africa and are distinct from those from Senegal collected in 2015 (MK940790 and MK940791).

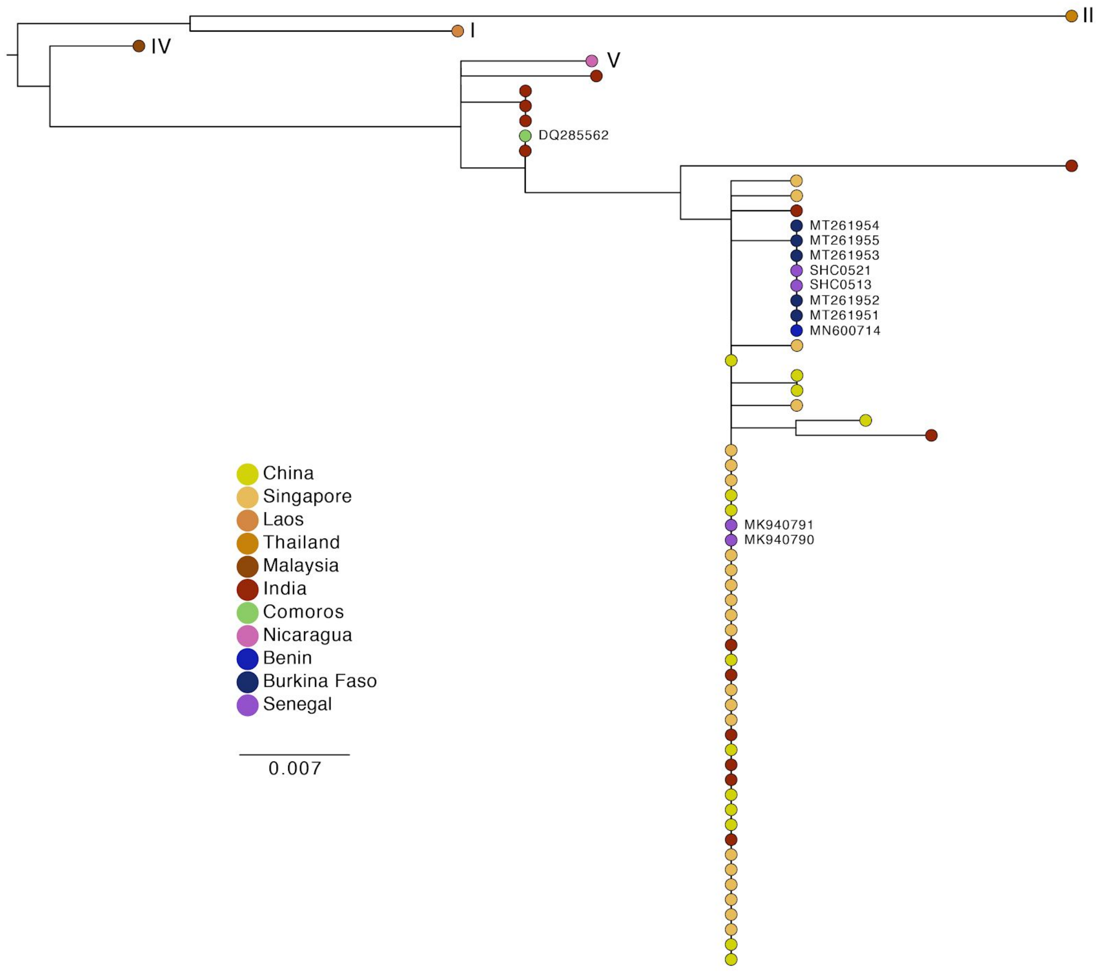

**Table S1.** Sequencing metrics for samples in the study in which dengue virus was detected by RT-qPCR or sequencing.

| Sample ID | Virus serotype | Total mapped reads pairs | Assembled genome length | CT mean |
|---|---|---|---|---|
| SHC0255 | DENV2 | 2832555 | 10659 | 33.04 |
| SHC0271 | DENV3 | 233966 | 10513 | 34.3 |
| SHC0406 | DENV3 | 821230 | 10525 | 35.12 |
| SHC0411 | DENV3 | 1436468 | 10655 | 32.85 |
| SHC0455 | DENV3 | 2022073 | 10680 | 28.91 |
| SHC0459 | DENV2 | 51926 | 10640 | 30.17 |
| SHC0479 | DENV2 | 1039515 | 10641 | 30.55 |
| SHC0503 | #N/A | #N/A | #N/A | 34.1 |
| SHC0506 | DENV3 | 2022819 | 10680 | 25.86 |
| SHC0513 | DENV1 | 2566696 | 10693 | #N/A |
| SHC0517 | DENV3 | 1816047 | 10650 | 24.63 |
| SHC0518 | DENV2 | 213657 | 10551 | 27.45 |
| SHC0520 | DENV3 | 412026 | 10641 | 24.66 |
| SHC0521 | DENV1 | 1999070 | 10683 | #N/A |
| SHC0522 | DENV3 | 1313920 | 10663 | 29.42 |
| SHC0531 | DENV3 | 602554 | 10639 | 28.2 |
| SHC0539 | DENV3 | 2754428 | 10662 | 23.6 |
| SHC0540 | #N/A | 2211767 | #N/A | 36.01 |
| SHC0543 | DENV3 | 713170 | 10650 | 26.9 |
